# Combination treatment of persistent COVID-19 in immunocompromised patients with remdesivir, nirmaltrevir/ritonavir and tixegavimab/cilgavimab

**DOI:** 10.1101/2023.04.07.23288144

**Authors:** Tal Brosh-Nissimov, Nir Ma’aravi, Daniel Leshin-Carmel, Yonatan Edel, Sharon Ben Barouch, Yafit Segman, Amos Cahan, Erez Barenboim

**Author notes:** **Corresponding author:** Dr. Tal Brosh-Nissimov, Head of the Infectious Diseases Unit, Samson Assuta Ashdod University Hospital, Harefua st. 7, Ashdod 7747629, Israel. **Author’s contributions** TBN - Conceptualization, formal analysis, writing – original draft. NM, DLC, YE, SBB, YS, AC, EB - Writing – review and editing.

## Abstract

**Background:** Little data exists to guide the treatment of persistent COVID-19 in immunocompromised patients. We have employed a unique protocol combining tixegavimab/cilgavimab, and short-term combination antivirals including remdesivir.

**Methods:** A retrospective single-center analysis of persistent COVID-19 in immunocompromised patients. Response was assessed by symptom resolution, declining C-reactive protein (CRP) levels and increasing SARS-CoV-2-PCR cycle-threshold (Ct) values.

**Results:** Fourteen patients were included, including 2 kidney transplant recipients, 11 with B-cell lymphoproliferative disease, treated with anti-CD20 or ibrutinib, and 1 with rheumatoid arthritis, treated with anti-CD20. Median Ct-value was 27 (interquartile range (IQR):24-32). All patients received tixegavimab/cilgavimab and a 5-day course of remdesivir. Eleven also received nirmaltrevir/ritonavir and one received molnupiravir. Median follow-up was 45 days (IQR:12-89). Eleven patients had complete responses including symptom resolution, decrease in CRP, and increase in Ct values (all with either a negative PCR or Ct value>30 on day 4-16). Three patients had a partial response with relapses requiring re-admission. One had died, and two responded to prolonged antiviral treatments.

**Conclusions:** A combination of monoclonal antibodies with antivirals has led to complete resolution of persistent COVID-19 in most severely-immunocompromised patients. Controlled studies will further direct the treatment of these patients, while more effective antivirals are urgently needed.

**Key points:** Some immunocompromised patients develop persistent symptomatic SARS-CoV-2 infection. Combination of monoclonal antibodies plus one or more antivirals cured 11/14 patients. Non-responders benefitted from prolonged combination antiviral treatment. Controlled trials are needed to find optimal treatment of persistent COVID-19.

## Introduction

The COVID-19 pandemic has become more manageable, due to highly efficient vaccines, development of immunity from natural infections, and antiviral drugs. Nevertheless, severely immunocompromised patients may have diminished vaccine efficacy and might be unable to clear severe-acute-respiratory-syndrome-coronavirus-2 (SARS-CoV-2) infection [1]. Patients with hematological malignancies, recipients of B-cell depleting therapy (BCDT) and transplanted patients can develop persistent COVID-19, defined as prolonged symptomatic infection, with continuing SARS-CoV-2 replication, and lack of serologic responses [2]. Patients with this syndrome frequently relapse shortly after treatment, resulting in significant morbidity and mortality [3]. The optimal treatment of persistent COVID-19 remains unknown. Challenged by many persistent COVID-19 patients during the Omicron-variant wave in Israel, we empirically implemented a treatment protocol combining monoclonal antibodies, antivirals, and corticosteroids. This study retrospectively analyses the treatment of persistent COVID-19 patients and their clinical course in the Samson Assuta Ashdod University Hospital.

## Materials and Methods

Beginning in March 2022, our treatment protocol for COVID-19 in highly immunocompromised patients was a combination of tixegavimab/cilgavimab, antivirals (remdesivir, nirmaltrevir/ritonavir, molnupiravir or a combination of any), and corticosteroids. tixegavimab/cilgavimab and combination antivirals were given with an off-label indication, after approval by the local pharmacy committee according to the Israeli Ministry of health regulations. Data was collected retrospectively and summarized. Viral replication status was estimated by the cycle threshold (Ct) value of SARS-CoV-2 PCR tests, with lower values representing higher viral loads. Anti-S antibody levels were measured by the Liaison® (DiaSorin, Saluggia, Italy) SARS-CoV-2 Trimeric IgG assay (negative, ≤12u/ml, borderline 13-22u/ml, positive >22u/ml, linearity range up to 800u/ml). Patients were followed during their hospital stay, and in some cases visited our outpatient clinic after discharge. Response to treatment was evaluated by symptom improvement, decrease in C-reactive protein (CRP) levels and negative PCR or increasing Ct values. The infecting SARS-CoV-2 variant was categorized as definite when identified with sequencing, or probable according to the most common (>50% frequency) circulating variant in Israel at the onset symptoms [4].

## Results

### Patients’ baseline characteristics

Fourteen immunosuppressed patients (9 males) received combination treatment (see table 1). The median age was 74 years (interquartile range, IQR, 69,79). Two patients were kidney transplant recipients on immunosuppressive treatment. Eleven patients had a B-cell lymphoproliferative disease: Seven patients had non-Hodgkin B-cell lymphoma (4 follicular, 1 mantle-cell, 1 mucosal-associated lymphoid tissue (MALT) and 1 marginal-zone histological types), and 4 with chronic lymphocytic leukemia (CLL). All the lymphoma patients received BCDT, a median of 165 days (IQR 46,704) before disease onset. Three CLL patients were treated with ibrutinib. One patient had rheumatoid arthritis, treated with BCDT. Comorbidities associated with COVID-19 included hypertension (8 patients), diabetes mellitus (6), chronic renal failure (4), ischemic heart disease (3), congestive heart failure (3), chronic lung disease (3), obesity (2), and chronic liver disease in 1 patient. Nine patients had received 4 vaccine doses against COVID-19, a median of 146 (IQR 119,244) days before disease onset, which was considered adequate according to Israeli recommendations. Five patients were not adequately vaccinated: 1 received 3 doses 262 days before disease onset, 2 received 2 doses more than a year before disease onset, and 2 were unvaccinated. Only three patients had received tixegavimab/cilgavimab prophylaxis 86, 104 and 114 days before disease onset, all with the standard 150/150mg dose that was used in Israel until September 2022.

**Table 1:**
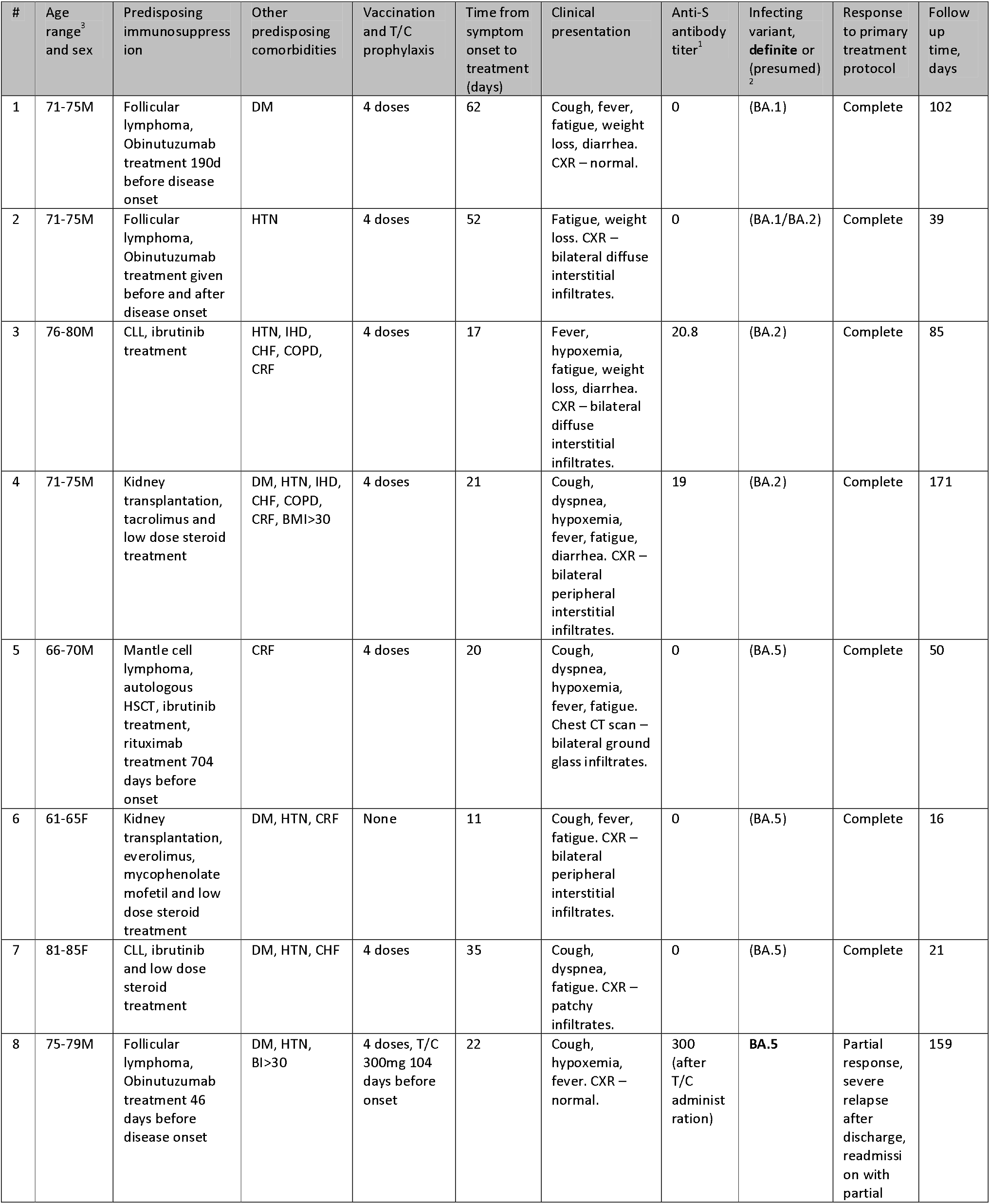
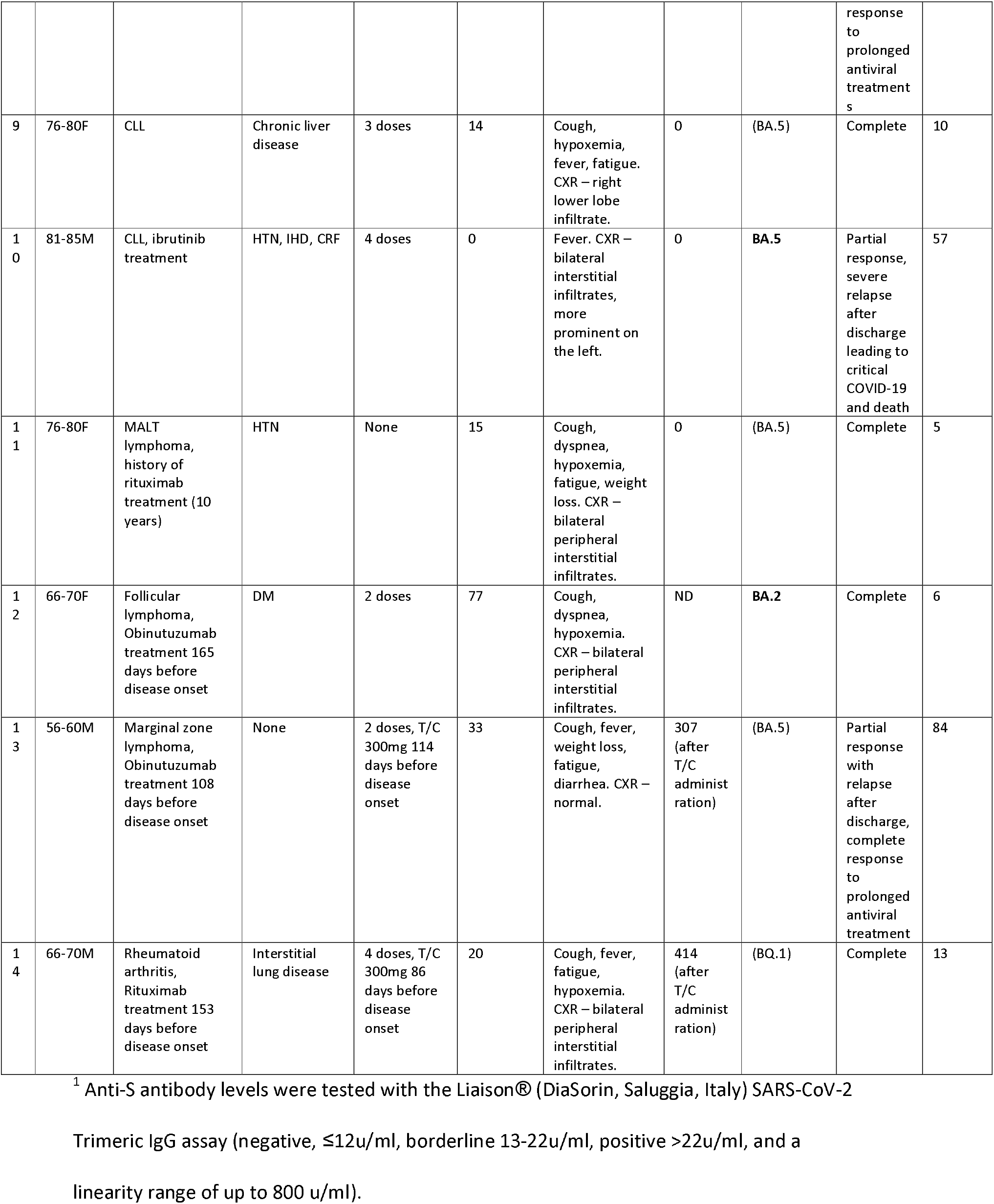
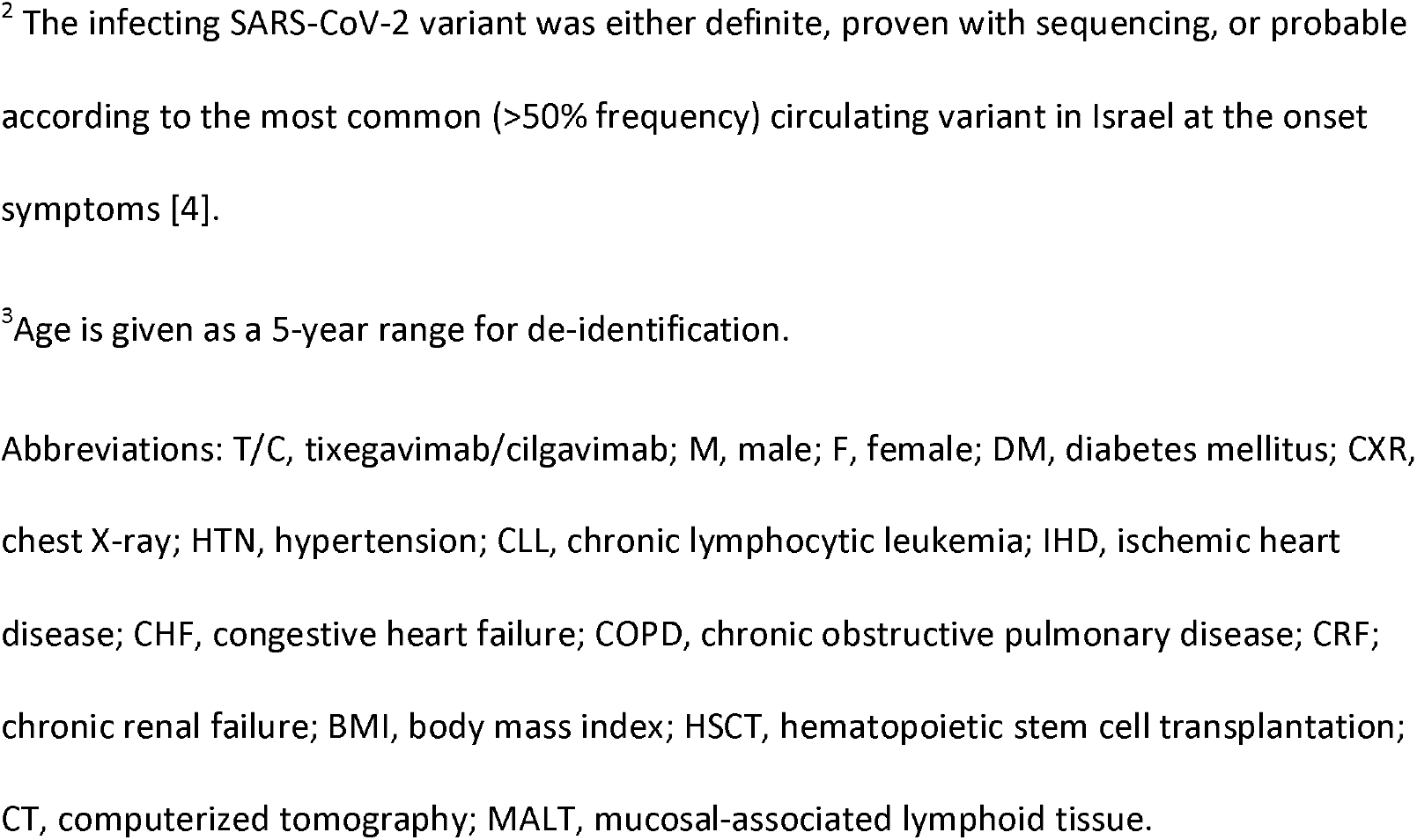
Clinical characteristics, treatments, and responses of persistent COVID-19 in immunocompromised patients

| # | Age range<sup>3</sup> and sex | Predisposing immunosuppression | Other predisposing comorbidities | Vaccination and T/C prophylaxis | Time from symptom onset to treatment (days) | Clinical presentation | Anti-S antibody titer<sup>1</sup> | Infecting variant, definite or (presumed)<sup>2</sup> | Response to primary treatment protocol | Follow up time, days |
| --- | --- | --- | --- | --- | --- | --- | --- | --- | --- | --- |
| 1 | 71-75M | Follicular lymphoma, Obinutuzumab treatment 190d before disease onset | DM | 4 doses | 62 | Cough, fever, fatigue, weight loss, diarrhea. CXR – normal. | 0 | (BA. 1) | Complete | 102 |
| 2 | 71-75M | Follicular lymphoma, Obinutuzumab treatment given before and after disease onset | HTN | 4 doses | 52 | Fatigue, weight loss. CXR – bilateral diffuse interstitial infiltrates. | 0 | (BA. 1/BA.2) | Complete | 39 |
| 3 | 76-80M | CLL, ibrutinib treatment | HTN, IHD, CHF, COPD, CRF | 4 doses | 17 | Fever, hypoxemia, fatigue, weight loss, diarrhea. CXR – bilateral diffuse interstitial infiltrates. | 20.8 | (BA. 2) | Complete | 85 |
| 4 | 71-75M | Kidney transplantation, tacrolimus and low dose steroid treatment | DM, HTN, IHD, CHF, COPD, CRF, BMI>30 | 4 doses | 21 | Cough, dyspnea, hypoxemia, fever, fatigue, diarrhea. CXR – bilateral peripheral interstitial infiltrates. | 19 | (BA. 2) | Complete | 171 |
| 5 | 66-70M | Mantle cell lymphoma, autologous HSCT, ibrutinib treatment, rituximab treatment 704 days before onset | CRF | 4 doses | 20 | Cough, dyspnea, hypoxemia, fever, fatigue. Chest CT scan – bilateral ground glass infiltrates. | 0 | (BA. 5) | Complete | 50 |
| 6 | 61-65F | Kidney transplantation, everolimus, mycophenolate mofetil and low dose steroid treatment | DM, HTN, CRF | None | 11 | Cough, fever, fatigue. CXR – bilateral peripheral interstitial infiltrates. | 0 | (BA. 5) | Complete | 16 |
| 7 | 81-85F | CLL, ibrutinib and low dose steroid treatment | DM, HTN, CHF | 4 doses | 35 | Cough, dyspnea, fatigue. CXR – patchy infiltrates. | 0 | (BA. 5) | Complete | 21 |
| 8 | 75-79M | Follicular lymphoma, Obinutuzumab treatment 46 days before disease onset | DM, HTN, BI>30 | 4 doses, T/C 300mg 104 days before onset | 22 | Cough, hypoxemia, fever. CXR – normal. | 300 (after T/C administration) | BA.5 | Partial response, severe relapse after discharge, readmission with partial response to prolonged antiviral treatments | 159 |
| 9 | 76-80F | CLL | Chronic liver disease | 3 doses | 14 | Cough, hypoxemia, fever, fatigue. CXR – right lower lobe infiltrate. | 0 | (BA.5) | Complete | 10 |
| 10 | 81-85M | CLL, ibrutinib treatment | HTN, IHD, CRF | 4 doses | 0 | Fever. CXR – bilateral interstitial infiltrates, more prominent on the left. | 0 | **BA.5** | Partial response, severe relapse after discharge leading to critical COVID-19 and death | 57 |
| 11 | 76-80F | MALT lymphoma, history of rituximab treatment (10 years) | HTN | None | 15 | Cough, dyspnea, hypoxemia, fatigue, weight loss. CXR – bilateral peripheral interstitial infiltrates. | 0 | (BA.5) | Complete | 5 |
| 12 | 66-70F | Follicular lymphoma, Obinutuzumab treatment 165 days before disease onset | DM | 2 doses | 77 | Cough, dyspnea, hypoxemia. CXR – bilateral peripheral interstitial infiltrates. | ND | **BA.2** | Complete | 6 |
| 13 | 56-60M | Marginal zone lymphoma, Obinutuzumab treatment 108 days before disease onset | None | 2 doses, T/C 300mg 114 days before disease onset | 33 | Cough, fever, weight loss, fatigue, diarrhea. CXR – normal. | 307 (after T/C administration) | (BA.5) | Partial response with relapse after discharge, complete response to prolonged antiviral treatment | 84 |
| 14 | 66-70M | Rheumatoid arthritis, Rituximab treatment 153 days before disease onset | Interstitial lung disease | 4 doses, T/C 300mg 86 days before disease onset | 20 | Cough, fever, fatigue, hypoxemia. CXR – bilateral peripheral interstitial infiltrates. | 414 (after T/C administration) | (BQ.1) | Complete | 13 |

<sup>1</sup> Anti-S antibody levels were tested with the Liaison® (DiaSorin, Saluggia, Italy) SARS-CoV-2
Trimeric IgG assay (negative, ≤12u/ml, borderline 13-22u/ml, positive >22u/ml, and a linearity range of up to 800 u/ml).
<sup>2</sup> The infecting SARS-CoV-2 variant was either definite, proven with sequencing, or probable according to the most common (>50% frequency) circulating variant in Israel at the onset symptoms [4].
<sup>3</sup> Age is given as a 5-year range for de-identification.
Abbreviations: T/C, tixegavimab/cilgavimab; M, male; F, female; DM, diabetes mellitus; CXR, chest X-ray; HTN, hypertension; CLL, chronic lymphocytic leukemia; IHD, ischemic heart disease; CHF, congestive heart failure; COPD, chronic obstructive pulmonary disease; CRF; chronic renal failure; BMI, body mass index; HSCT, hematopoietic stem cell transplantation; CT, computerized tomography; MALT, mucosal-associated lymphoid tissue.

On presentation, among patients who had not received tixegavimab/cilgavimab prophylaxis, anti-S antibody was negative in eight patients and borderline in 2. The infecting SARS-Cov-2 variant was definite in 3 patients, and probable in the rest, with 1, 4, 8 and 1 cases with BA.1, BA.2, BA.5, and BQ.1/BQ.1.1, respectively.

### Clinical presentation

The median time from disease onset to presentation was 20 (IQR 15,39). All patients sought care due to significant symptoms, either present continuously from disease onset, or recurring after a short phase of clinical improvement. Prominent symptoms included cough (11 patients), fatigue (11), fever (10), dyspnea (5), weight loss (4) and diarrhea (4). Nine patients were hypoxemic. In many cases of prolonged symptoms, patients had been treated elsewhere for respiratory tract infections with recurrent courses of antimicrobials. Most patients had lymphopenia on presentation, with a median count of 500/ml (IQR 300,700). CRP was elevated in all cases, with a median value of 107 (IQR 67,130) mg/L. Twelve patients had abnormal findings in chest x-ray or CT scan, mainly diffuse or patchy bilateral interstitial infiltrates. All patients had a positive PCR result for SARS-CoV-2, with a median Ct value on presentation of 27 (IQR 24,32). Ten and five patients had Ct value below 30 and 25, respectively.

### Treatment

All patients received intramuscular tixegavimab/cilgavimab. Dosing of tixegavimab/cilgavimab varied: Ten patients were administered 300/300mg according to the dosing in the TACKLE study [5], and 4 patients were administered 150/150mg, including two who had received prophylactic tixegavimab/cilgavimab before disease onset. All patients received a 5-day course of intravenous remdesivir (200mg loading dose followed by 100mg/d). Eleven patients received nirmaltrevir/ritonavir 300/100mg bid for five days. Two transplanted patients (#4,6) had a major drug-drug interaction precluding ritonavir administration. One was administered molnupiravir 800mg bid for five days, and the other received single-agent treatment with remdesivir. One lymphoma patient refused nirmaltrevir/ritonavir treatment. Twelve patients received dexamethasone 6mg daily for 5-10 days.

### Response to treatment

Median follow-up was 45 (IQR 12,89) days. All 14 patients had a subjective symptomatic improvement at the end of treatment. Three patients (#8,10,13) were readmitted due to clinical relapse with either fever or dyspnea occurring soon after discharge. These three patients will be referred to as partial responders and will be discussed separately. Of eleven patients with a complete response, PCR tests taken between day 4-16 from treatment onset were negative in 7 cases, and with a Ct value >30 in 4 additional cases (see Figure 1). CRP levels gradually decreased in all eleven complete responders (see Figure 2).

**Figure 1:**
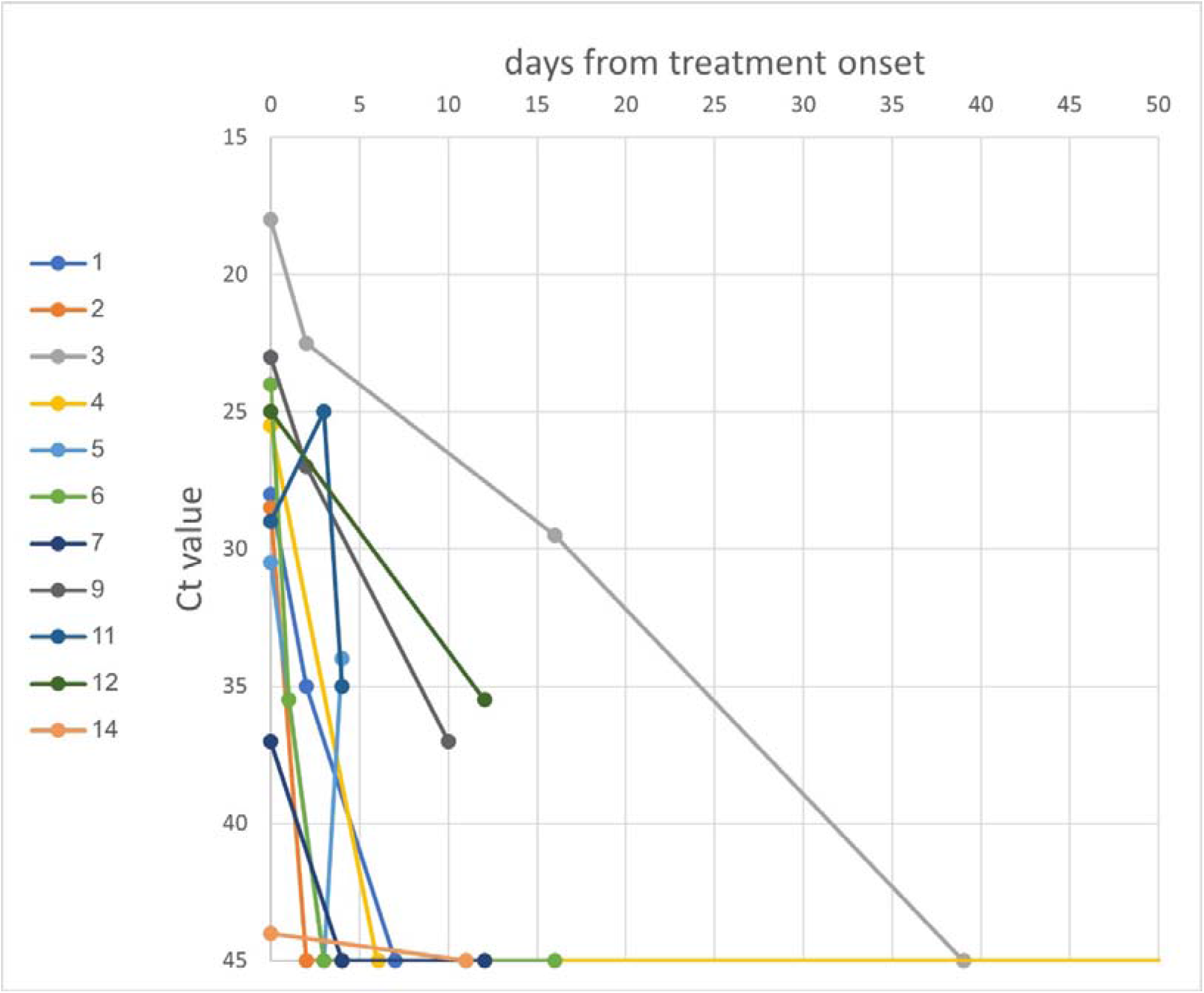
SARS-CoV-2 viral burden in patients who had a sustained response to combination treatment. Colors represent individual patients (see Table 1). Higher Ct value represent lower viral RNA load. A Ct value of 45 signifies a negative PCR.

**Figure 2:**
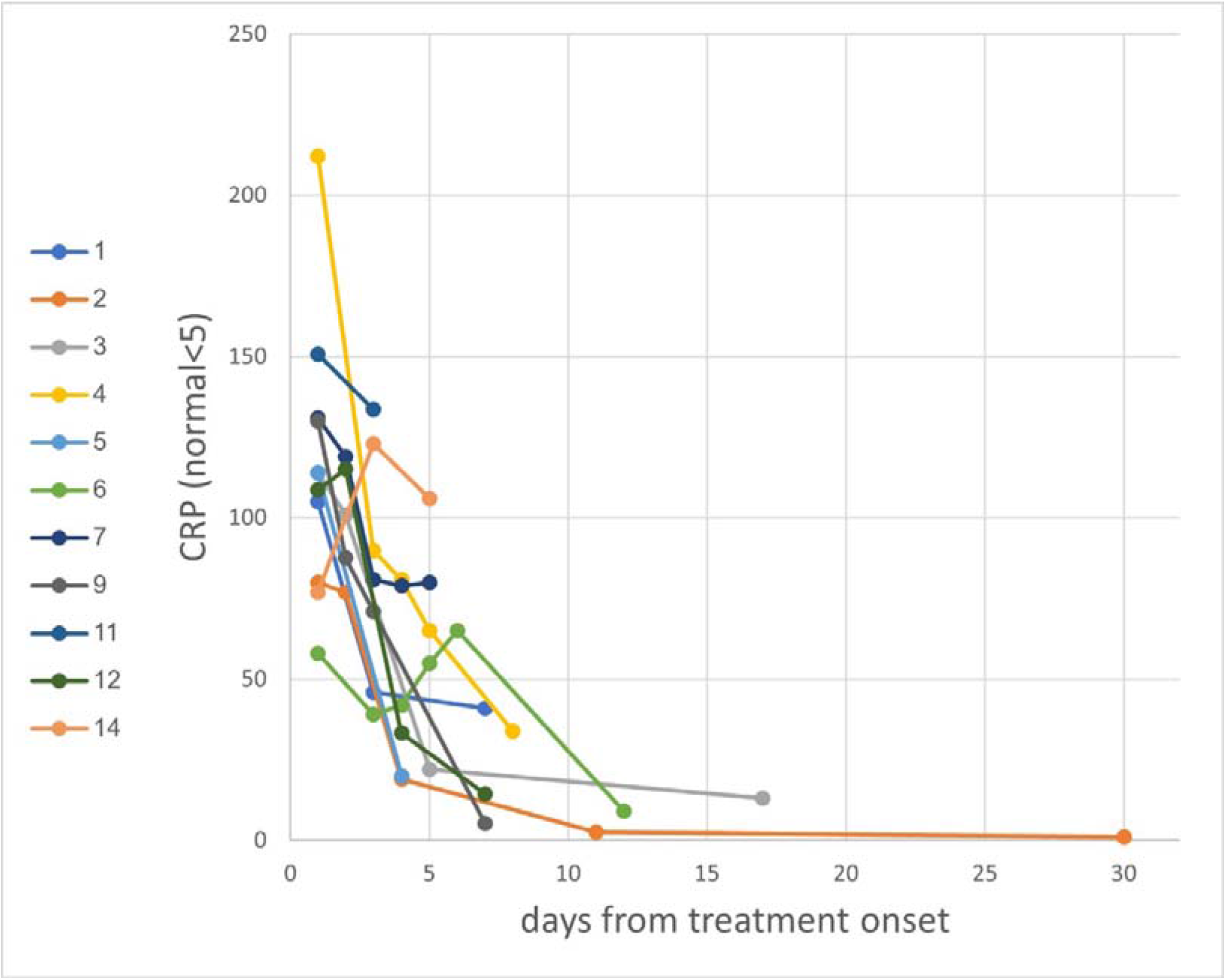
C-reactive protein (CRP) levels in patients who had a sustained response to combination treatment.

### Description of partial responders (see also figure 3)

Patient 10: A male, 81-85 years of age, with CLL treated with frontline ibrutinib. The patient presented with fever and pulmonary infiltrates on chest X-ray. SARS-CoV-2 sequencing was compatible with the BA.5 variant. After 5 days of treatment with remdesivir, nirmaltrevir/ritonavir and tixegavimab/cilgavimab 300/300mg, he was discharged without fever. Five weeks later the patient presented with recurrent fever for almost 4 weeks, cough, dyspnea, and hypoxemia. He was treated with broad-spectrum antibiotics, steroids, 14-day course of baricitinib and two short courses of remdesivir. He developed respiratory failure, for which he was mechanically ventilated, and died after 22 days.

**Figure 3:**
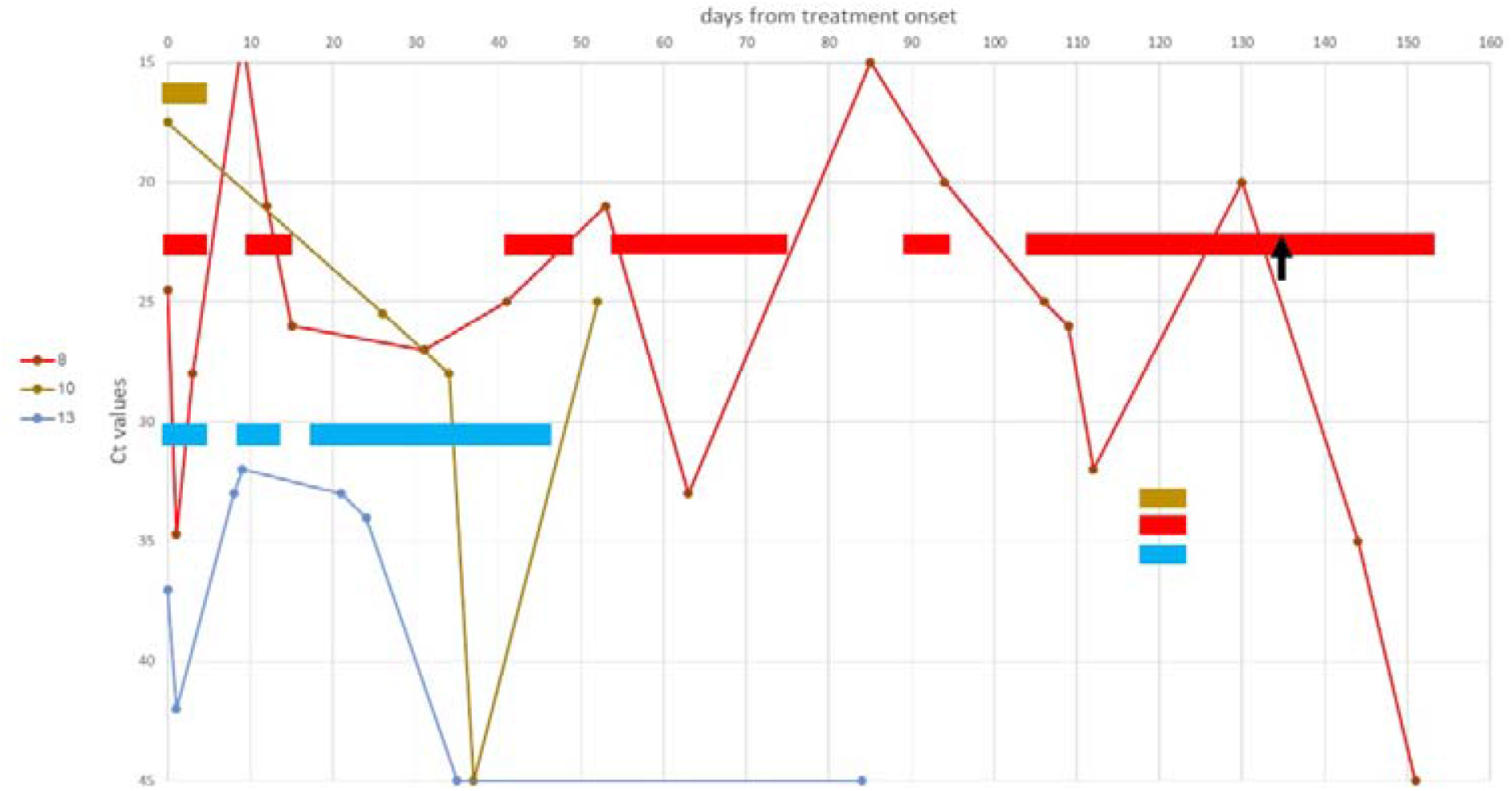
SARS-CoV-2 PCR cycle threshold (Ct) values in three patients who had a partial response to combination treatment, with relapsed after discharge. Colors represent individual patients (see Table 1). Rectangular blocks signify periods of antiviral treatments. Higher Ct values represent lower viral RNA load. A Ct value of 45 signifies a negative PCR. The arrow marks the addition of molnupiravir to nirmaltrevir/ritonavir monotherapy.

Patient 13: A male, 56-60 years of age, with a history of marginal-zone lymphoma, treated with bendamustine and rituximab. The last dose of rituximab was given 108 days before disease onset. He was diagnosed with mild COVID-19 and treated as an outpatient with nirmaltrevir/ritonavir for 5 days. He continued to suffer from fever, fatigue, cough, diarrhea, and a lost 10Kg in weight. No viral sequencing was done, but his symptoms began during a BA.5-predominant period. After one month of symptoms and a few antibiotic treatments, he was admitted and treated with remdesivir for 5 days, tixegavimab/cilgavimab 150/150mg, and dexamethasone, but refused nirmaltrevir/ritonavir treatment. Four days after discharge the patient was readmitted with symptom recurrence, and was treated with remdesivir for 5 days, tixegavimab/cilgavimab 150/150mg, dexamethasone and baricitinib. Eight days after his second admission the patient was readmitted with the same symptoms. On his third admission, a combination of remdesivir and nirmaltrevir/ritonavir, both were given and continued after discharge for a completion of 14 days (remdesivir) and 30 days (nirmaltrevir/ritonavir). During this treatment and for >2 months after the end of treatment, the patient had complete resolution of his symptoms and negative PCR tests.

Patient 8: A male, 71-75 year of age, with a history of follicular lymphoma, treated with bendamustine and obinutuzumab (last dose was given 46 days before disease onset). He was admitted 22 days after diagnosis of COVID-19, due to persistent cough, fever, and hypoxemia, and treated with remdesivir and nirmaltrevir/ritonavir for 5 days, tixegavimab/cilgavimab 150/150mg and dexamethasone. SARS-CoV-2 sequencing identified the BA.5 variant. Following that admission, He was readmitted twice with progressive cough, dyspnea, hypoxemia, and received a few courses of antibiotics, corticosteroids and remdesivir. On his fourth admission significant respiratory deterioration was noted. More corticosteroids and a course of baricitinib was given, without clinical improvement. A lymphocyte populations count showed a CD4 count of 20 cells/μL. Bronchoalveolar lavage specimens were positive for SARS-CoV-2, *Mycobacterium simiae* and cytomegalovirus (CMV), and blood PCR yielded a CMV-RNA’emia of 352,000 copies/ml. Treatment with ganciclovir and antimycobacterial drugs was started. During this admission remdesivir and nirmaltrevir/ritonavir were given for 20 days. During this course, clinical improvement, and an increase of Ct values from 21 to 33 were noted, but in a repeat PCR after treatment, a Ct value of 15 was found. The patient was put on another prolonged combined treatment with significant improvement enabling discharge. Nirmaltrevir/ritonavir monotherapy was continued for three weeks, with decreasing Ct values. SARS-CoV-2 PCR became negative after the addition of molnupiravir. The patient still receives antiviral treatment at home.

## Discussion

In this retrospective analysis of fourteen immunocompromised patients with or at-risk for persistent COVID-19, combination treatment including two antiviral medications and monoclonal antibodies, led to complete resolution of clinical and virological evidence of infection in 11 patients, and partial responses in three.

A functional adaptive B-cell response is important in SARS-CoV-2 clearance [1]. Patients receiving BCDT are at an elevated risk for severe COVID-19 and mortality. In a large cohort study, 43.9% of COVID-19 patients who received BCDT required hospitalization, 8.8% have died, and 17.3% relapsed [3]. Similar outcomes were also described during the Omicron variant period [6–8]. Increased risk for COVID-19 persistence in immunocompromised patients was shown with anti-CD20 therapy and lower CD4 lymphocyte counts [9]. These patients commonly present with fever, cough, dyspnea, radiological signs of pneumonitis, and positive SARS-CoV-2 PCR tests, lasting for a prolonged period, sometimes in a remitting-relapsing pattern, while no serologic response develops. Definitions for persistent and seronegative COVID-19 were suggested by Belkin et al [2], to include symptom duration of 7-14 days and PCR positivity of >21 days. Most of our patients fulfilled these definitions, although some had a shorter disease duration.

Contemporary treatment of COVID-19 includes antiviral medications, monoclonal antibodies and various immunomodulators, all of which have been proven beneficial in high-quality clinical trials. However, severely immunocompromised patients were excluded from most of these. Due to limited evidence, clinical guidelines such as those published by the National Institute of Health (NIH), do not recommend a different approach to the treatment of immunocompromised patients with COVID-19 [10,11]. Nevertheless, as noted, COVID-19 in immunocompromised patients tends to have a different course and thus, a different approach may be warranted. Due to the inability to mount an adaptive serological response and the prolonged SARS-CoV-2 replication in these patients, administering antivirals and antibody treatments beyond the first days of disease and continuing these agents for a longer duration than used in clinical trials have at least theoretical justification. At the same time, the use of immunomodulators such as steroids, baricitinib and anti-IL6 agents might carry a greater risk of secondary infections in these already immunodeficient patients. In order to maximize the chances of success, while limiting the risks for antiviral resistance that was reported in persistent COVID-19 patients receiving remdesivir [12], we have chosen to combine, whenever possible, two antiviral medications, in addition to tixegavimab/cilgavimab, a monoclonal antibody preparation that was available in Israel with a proven neutralizing activity against the circulating Omicron variants.

Evidence on successful treatment of immunocompromised patients with persistent COVID-19 is limited to retrospective case series and cohort studies. The earliest successful reports were for convalescent plasma therapy (CPT) [13]. With the availability of antivirals and monoclonal antibodies, concurrent and sequential treatments were reported. Combined remdesivir and CPT were successfully used shortly after COVID-19 symptom onset, in 20 patients with hematological malignancies and severe lymphopenia [14]. Earlier treatment and simultaneous administration of the antiviral with CPT were associated with more rapid resolution of supplemental oxygen dependency and with shorter hospital stay. In a study evaluating the outcomes of various treatments in 31 cases of persistent COVID-19, mostly after BCDT, a combination of remdesivir and either CPT or casirivimab/imdevimab was associated with better chances of symptomatic improvement and viral clearance than remdesivir only [15]. In a few reports, monoclonal antibodies were curative after a very prolonged infection, sometimes unresponsive to recurrent treatments with remdesivir and steroids [16–21]. Combination treatment of antivirals and monoclonal antibodies, usually with steroids, was also reported successful in persistent COVID-19 [17,22–24].

Nevertheless, three of our patients did not achieve complete and sustained response after a 5-day antiviral treatment and tixegavimab/cilgavimab. All three had a BA.5 infection (two definite and one probable), while 6/11 responders were infected by BA.5 or BQ.1, which are not neutralized by tixegavimab/cilgavimab as efficiently as previous Omicron variants [25]. This difference was not statistically significant (χ^2^ test, *p*=0.26), as were other possible differences between responders and non-responders, including age, comorbidities, and lymphocyte counts (data not shown). One of the non-responders (#13) refused nirmaltrevir/ritonavir during the primary treatment course and was treated with remdesivir and tixegavimab/cilgavimab only. With the first non-responding patient (#10), upon presentation with progressive COVID-19 pneumonitis, we used the recommended immunomodulatory treatments including steroids and baricitinib, and the patient had subsequently deteriorated and died. Another non-responder also did not respond to this approach but had a sustained full recovery with a prolonged combination antiviral treatment (14 days of remdesivir, 30 days of nirmaltrevir/ritonavir). The third non-responder (#8) developed significant opportunistic infections after steroids and baricitinib treatment, and subsequently achieved partial responses with combination antivirals. These three cases suggest that immunomodulatory treatment of severe COVID-19 in severely immunocompromised patients might carry more risks than benefit, and that prolonged antiviral treatments might be useful for sustained responses or even cure.

This study has a few important limitations. Patients were not treated using a systematic treatment and follow-up protocol. The treatment approach described was used for all immunocompromised patients beginning in March 2022, however, we could not use a historical control group for comparison, as other SARS-CoV2 variants were prevalent during the earlier period of the pandemic. As most patients received a combination of various treatments, we do not know whether responses were due to one of those treatments or their combination. We had sequencing results for only three patients, although the variants of the other patients could be presumed according to the national epidemiology. The follow-up time was short for some patients, but had these patients relapsed, it is likely they would have returned for care in our hospital, being the only hospital in town.

In summary, we show that severely immunocompromised patients with persistent COVID-19 can be successfully treated with a combination of antivirals and monoclonal antibodies, either with a short 5-day treatment, or, for non-responders, with repeated and prolonged antiviral treatments. Non-response might have been associated with the BA.5 variant, which was not as well-neutralized by tixegavimab/cilgavimab. This report, together with others, calls for controlled studies to further assess the optimal treatment for persistent COVID-19, and shows the importance of continuous development of new antivirals and variant-adapted monoclonal antibodies.

## Data Availability

All data produced in the present study are available upon reasonable request to the authors.

## Statements

### Ethical approval

The study was conducted according to the guidelines of the Declaration of Helsinki and was approved by the institutional review board (#AAA-22-113). Informed consent was waved due to the retrospective design.

